# Disentangling the effects of menopause on cognitive performance – causal framework and findings of the LIFE-Adult-Study

**DOI:** 10.1101/2025.03.06.25323411

**Authors:** Nadine Bonberg, Andrea E. Zülke, Susanne Röhr, Melanie Luppa, Kerstin Wirkner, Ronny Baber, Matthias L. Schroeter, Arno Villringer, Julia Sacher, Veronica Witte, Christoph Engel, Markus Löffler, Steffi Riedel-Heller, Heike Minnerup

## Abstract

**Objective:** To investigate the effects of age at natural menopause and reproductive lifespan on cognitive performance using causal reasoning.

**Methods:** We first identified potential mechanisms linking menopause and cognition focusing on the potential causal role of cumulative estrogen exposure. We then used cross-sectional data from approximately 2,200 postmenopausal women from the population-based LIFE-Adult Study to examine the effect of age at natural menopause and reproductive lifespan, as proxies for cumulative estrogen exposure, on neuropsychological test scores (Trail Making Tests A&B and CERAD Word Fluency Test), considering potential confounding and moderating variables. Results: Assuming that both age at menopause and reproductive lifespan are indicators of cumulative estrogen exposure, the data analysis revealed that women with later age at menopause and longer reproductive lifespan, respectively, performed better on all neuropsychological tests.

**Conclusion:** It is crucial to identify the causal framework when investigating the effects of women specific factors on cognition, especially when it comes to time-dependent effects, as well as complex moderating and mediating factors.

## 1 Introduction

Characterizing the effects of women-specific risk factors for cognitive aging is necessary to improve prevention and personalized treatment. Menopause, defined as the stage of a woman’s life when her menstrual periods stops permanently [1], is associated with cognitive decline beyond that expected with increasing age [2,3]. This is likely due to a complex interplay of hormonal, neurobiological, and psychosocial factors [4]. One of the major factors in this interplay is estradiol. Estradiol plays crucial roles in neurotransmitter regulation, synaptic plasticity, and neuroprotection [5–7]. However, the relationship between estrogen and cognition is complex, with two aspects of estrogen exposure in particular emerging as relevant for cognitive outcomes: first, the cumulative lifetime exposure, which may contribute to cognitive reserve [6,8] and which depends substantially on age at menopause; second, the dramatic decline in circulating estradiol levels during the menopausal transition [3,9]. These factors, individually or in combination, have been proposed as key mechanisms underlying cognitive changes associated with menopause, although findings are not always consistent and causal relationships remain to be definitively established [10]. A major challenge is to disentangle the cognitive effects of menopause from other negative cognitive effects associated with the menopausal transition that result from increasing age and an age-related increasing risk of cardiovascular disease, making it difficult to separate the cognitive effects specifically attributed to menopause from those related to the natural aging process. The effect of cardiovascular disease on menopause-related cognitive changes is particularly complex, as cardiovascular disease may also be a consequence of menopausal hormonal changes [11]. The relationship between menopause and cognition is also further complicated by several other factors, that may have confounding, moderating or mediating effects, including depression, cardiovascular disease, stress, and genetic susceptibility, such as the apolipoprotein E (APOE) allele status [12].

Given this complex interplay of factors, there is an urgent need to disentangle these effects and identify causal relationships to be able to inform targeted prevention strategies. The current study specifically aimed to identify the causal roles and effects of age at menopause and reproductive lifespan, as proxies for lifetime estrogen exposure, on cognition using causal inference methods. We first constructed Directed Acyclic Graphs (DAGs) [13] to represent our assumptions about the causal relationships and to identify the variables that should and should not be included in our statistical models. We then applied the models to a cross-sectional dataset of approximately 2,200 women aged 45-80 years from the population-based LIFE-Adult-Study to estimate the total effect of age at menopause and reproductive lifespan on several neuropsychological tests considering potential mediators, moderators, and confounders. This approach allows us to visually represent and analyze hypothesized causal pathways and apply them to real world data [14,15].

By addressing these questions through a causal inference framework, we aim to provide more robust evidence for the causal role of reproductive factors, particularly age at menopause, in cognitive aging. This approach may help clarify conflicting findings in the literature, inform targeted interventions for maintaining cognitive health in aging women, and improve the quality of research in this important area of women’s health.

## 2 Methods

### 2.1 Study population

The LIFE-Adult-Study is a population-based cohort study conducted by the Leipzig Research Center for Civilization Diseases (LIFE) in Leipzig, Germany, with a target population of 10,000 randomly selected participants [16]. The primary aim of the study is to investigate prevalences, genetic predispositions, early onset markers and the role of lifestyle-related factors for chronic lifestyle diseases. The study was approved by the responsible institutional ethics board of the Medical Faculty of the University of Leipzig (approval numbers: 263–2009-14122009, 263/09- ff, 201/17-ek). Written informed consent was obtained from all participants. The participation rate was 33% [16]. Baseline examinations were performed between 2011 and 2014.

All study participants underwent a core assessment program, including computer-assisted in-person interviews, computer- or paper-based self-administered questionnaires, physical and medical examinations, and neuropsychological testing. All assessments were conducted by trained study staff [16–20].

For the present analysis, we applied some exclusion criteria. We excluded participants with limited German language skills and severe neurological disorders. We also defined test-specific exclusion criteria, depending on the underlying neuropsychological test, and we included only women aged 45 years and older (see Figure 1).

**Figure 1:**
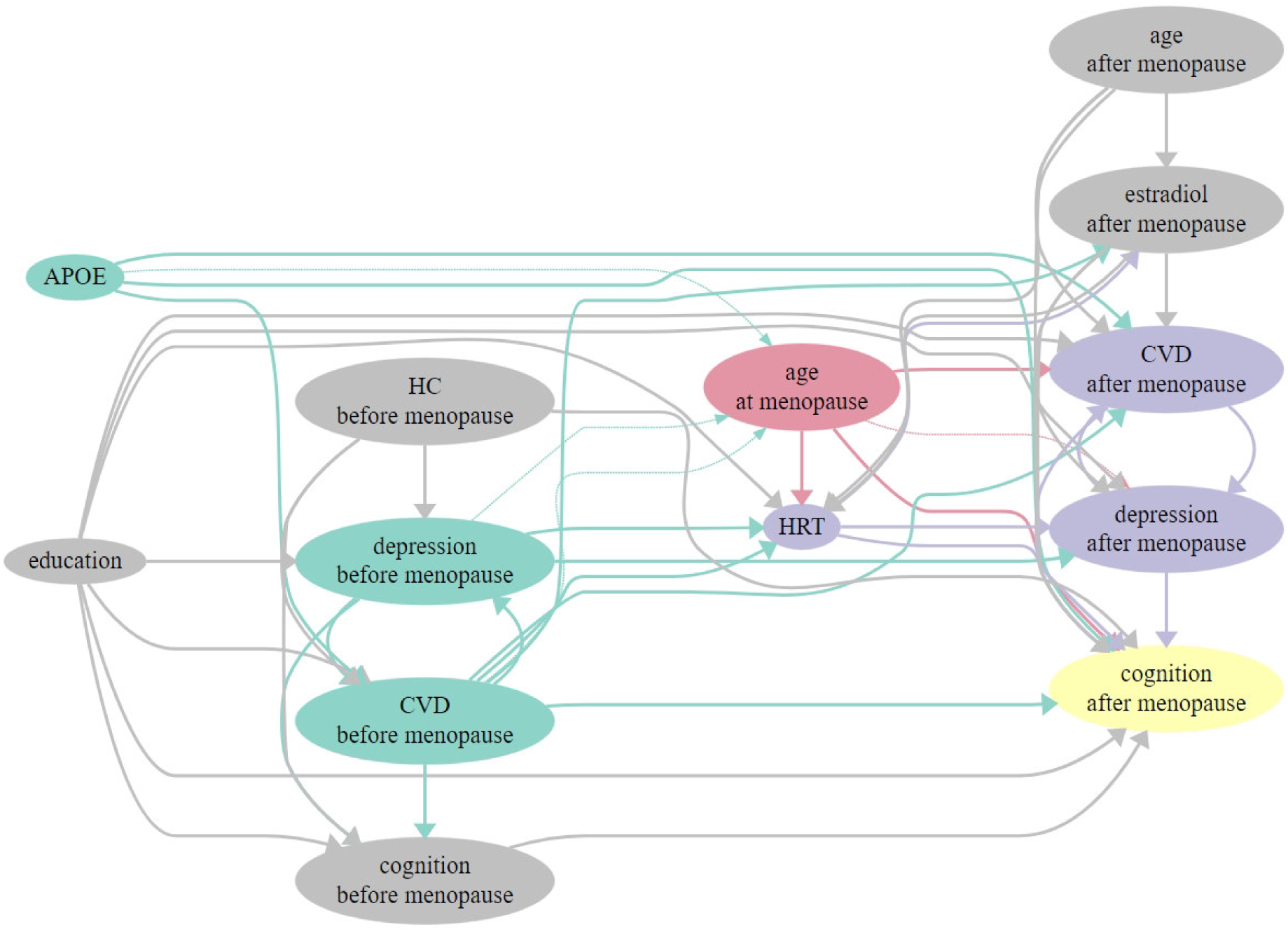
Directed acyclic graph (DAG) representing the hypothesized causal relationship between age at menopause (exposure) and cognition (outcome). Red: exposure, yellow: outcome, green: ancestors of exposure and outcome, grey: ancestors of outcome, purple: potential mediator of the exposure-outcome relationship. Abbreviations: HC: hormonal contraception; HRT: hormone replacement therapy, CVD: cardiovascular disease. To explore the hypothesized causal relationship between reproductive lifespan (exposure) and cognition (outcome), age at menopause is substituted for reproductive lifespan.

### 2.2 Exposure assessment

Data on menstrual cycle (time since last menstrual period (LMP)), history of bilateral oophorectomy and hysterectomy, and use of hormone replacement therapy were collected using a structured interview. Women were classified as postmenopausal if LMP was > 12 months or age was ≥ 55 years. Women aged ≥ 55 years and with LMP < 12 months were excluded from the analysis. In addition, women with a history of bilateral oophorectomy or hysterectomy were excluded from all analyses. Reproductive lifespan was defined as the time from menarche to the last menstrual period.

### 2.3 Covariate Assessment

#### 2.3.1 Sociodemographic characteristics and health status

Sociodemographic characteristics, education, smoking status, and data on participants’ health status and history were assessed in the core assessment program. Education was documented according to the socioeconomic status (SES)-index and categorized as low, middle, or high according to the Comparative Analysis of Social Mobility in Industrial Nations/CASMIN classification [21]. Body weight and height were measured in a standardized manner [16], and the body mass index (BMI) was calculated in kg/m^2^ and categorized as non-obese (BMI <30 kg/m^2^) and obese (BMI ≥ 30 kg/m^2^). Arterial hypertension was defined as a reported history of hypertension or use of antihypertensive medication, respectively. Diabetes was defined as a reported history of diabetes mellitus, use of antidiabetic medication, HbA1c levels >=6.6%, or blood glucose levels (oral glucose tolerance test (oGTT) 120 min) of >=11 mmol/l. Smoking status was defined categorically as never vs. former vs. current smoker. A cardiovascular risk score was calculated as the sum of the risk factors diabetes, current smoking, hypertension and obesity. For analysis, the score was divided into categories of 0-2 and 3-4 risk points. Depressive symptoms were identified using the Center for Epidemiological Studies Depression Scale (CES-D), a 20-item self-report screening instrument [22]. The CES-D score can reach a maximum of 60. Higher CES-D scores indicate more depressive symptoms. A CES-D score ≥ 16 has been used to define clinically relevant depressive symptoms [22]. The use of hormone replacement therapy was documented as never/ever.

#### 2.3.2 APOE genotype

Genomic DNA was isolated from peripheral blood leukocytes by staff of the Leipzig Medical Biobank using an automated protocol with the Qiagen Autopure LS (Qiagen, Hilden, Germany) to determine the APOE genotype. A NanoDrop spectrophotometer was used to specify DNA purity and yield. Subsequently, the APOE allele status (e2, e3, e4) was determined using a Roche Lightcylcer 480 according to the method of Aslanidis and Schmitz (1999) [23]. For statistical analyses, APOE e4 allele status was divided into two categories: (1) at least one e4 allele and (2) no e4 allele.

### 2.4 Outcome assessment

Validated neuropsychological tests were administered to the study participants. Participants completed the following tests:

*Trail-Making Test A&B* [24]: Participants were asked to connect consecutive numbers from 1 to 25 to measure attention and psychomotor speed (TMT A), and consecutive numbers and letters in an alternating sequence to measure mental set shifting (TMT B). The time taken to complete each part was recorded. The time limit to complete the TMT A was 180 s and 300 s for the TMT B.

*Verbal Fluency Test ‘Animals’* [25–27]: Participants were asked to verbally produce as many animal names as possible within 60 seconds.

The Trail-Making Test and the Verbal Fluency Test are tests of the extended *Consortium to Establish a Registry for Alzheimer’s Disease Neuropsychological Assessment Battery* (CERAD-NAB [25,26,28]).

TMT A and B scores showed a skewed distribution and were log-transformed prior to z-score calculation. Z-scores were calculated for all neuropsychological tests using the respective test mean and standard deviation (SD) of the female participants for standardization. All z-scores are scaled so that higher values represent better outcomes.

### 2.5 Directed Acyclic Graph (DAG)

We examined how age at menopause (exposure) influences cognition (outcome). To identify causal and biasing pathways, we used Directed Acyclic Graphs (DAGs). DAGs help determine the necessary adjustment variables for the analysis. When examining the causal effect of exposure on outcome, one can focus on either total or direct effects. The key difference lies in the treatment of mediators. Our study focused on total effects, which, unlike direct effects, do not require adjustment for mediators, but rather represent the “total”, i.e., the sum of all direct and indirect (mediating) effects of the exposure on the outcome.

We used the R package dagitty [29], to determine minimal sufficient adjustment sets (MSAS) to analyze the total effect of age at menopause on cognition and plotted the DAG using DiagrammeR version 1.0.11. [30]. Our decisions about variable inclusion, arrow placement, and directionality were based on an extensive literature review (Supplementary Table 1). In cases of uncertainty, we developed an additional adjustment set for use in sensitivity analysis. The DAG illustrating the effect of age at menopause on cognition is shown in Figure 1.

In the DAG, the total effect of age at menopause on cognition may comprise several direct and indirect effects: the direct effect would be the effect of age at menopause as indicator of cumulative estrogen exposure. Indirect effects would be effects mediated by e.g. use of HRT, that would be prescribed due to a woman’s specific age at menopause. For variables to act as confounders in the relationship between cumulative estrogen exposure / age at menopause and cognition they must be causes of both, age at menopause and cognition. Factors that might act as confounders are cardiovascular disease and depression. Our dataset uses a cardiovascular risk score as a proxy for cardiovascular disease (CVD) and the CES-D score as an indicator of current depressive symptoms. However, these measures are available only once, at the time of study participation, leaving us without information on their presence prior to menopause in this postmenopausal study population. This lack of longitudinal information complicates causal interpretation, as the timing of depression or CVD onset relative to menopause is crucial to understanding their role in the menopause-cognition relationship. If present before menopause, these conditions could potentially influence both the age at menopause and post-menopausal cognition, thus acting as confounders. However, it should be noted that the evidence for such confounding effects is currently weak (see Supplemental Table 1). Conversely, if these conditions developed post-menopause, they might serve as mediators in the causal pathway between menopause and cognition. Ideally, we would have both pre- and post-menopausal data on CVD and depression. This comprehensive data would allow us to account for the possibility that these factors may have both confounding and mediating effects, depending on their time of onset. Unfortunately, our data do not allow us to determine this temporal sequence. To address this limitation, we use a dual analytic approach. We conduct our analyses both with and without adjustment for the cardiovascular risk score and CES-D. In cases where we do adjust, we use the postmenopausal measures as proxies for the unobserved premenopausal measures. This strategy allows us to compare results under different assumptions about the temporal relationships between variables, while using the available postmenopausal data to approximate premenopausal conditions. The uncertainty regarding the temporal sequence is also reflected in the double-headed arrow (technically not allowed) in the DAG between depression and CVD, which can mutually influence each other [31]. Another potential confounder is APOE genotype. While the effect of APOE genotype on cognition is well known, there are some conflicting reports of an association between APOE genotype and age at menopause reporting that carriers of the APOE4 allele reach menopause at an earlier age [32] or later age [33]. Though this evidence is weak, we included APOE genotype in our MSAS. Alternatively, if not acting as a confounder, APOE genotype might be an effect moderator in the association between age at menopause and cognition [34]. We therefore performed sensitivity analyses including an interaction term of APOE genotype and age at menopause in our models.

Disentangling all these differential effects poses a significant challenge in observational studies, particularly cross-sectional ones. This complexity is further emphasized by the need to quantify lifetime estrogen exposure, which can only be approximated through various proxies. We chose to look at age at menopause. Another proxy is reproductive lifespan, which can serve as an alternative to age at menopause. To explore this approach, we conducted additional analyses using our original set of variables but substituted age at menopause for reproductive lifespan. More comprehensive methods involve constructing estrogen exposure scores, as demonstrated by Smith et al. [35], Rasgon et al. [36] or Fox et al. [8]. While these approaches may provide a more precise reflection of cumulative lifetime estrogen exposure, they obscure the individual (causal) effects of each variable and thus the potential targets for intervention.

### 2.6 Statistical analysis

Linear regression models for the three standardized neuropsychological tests (Word Fluency Test and TMT A&B, respectively) were performed, applying the adjustment sets described above. All statistical analyses were performed with R 4.3.0 [37] and RStudio version 2023.03.0+386 [38]. Analyses were performed with a 2-tailed alpha of 0.05 referred as the statistically significant level. For interactions, the significance level was set at 0.1.

To estimate the total effect of age at menopause on cognitive performance, we used different linear regression models with the cognitive test result as dependent variable and age at menopause as independent variable. Additionally, we used two different adjustment sets derived by our DAG approach described above:

1. basic adjustment: age age^2^, education, APOE e4-allele status
2. basic adjustment + CES-D score and cardiovascular risk score

To account for possible mediating or moderating effects of HRT we also performed these analyses in women not taking HRT.

To investigate a possible moderating effect of APOE e4-allele status in the relationship between age at menopause and cognitive performance, the interaction between age at menopause and APOE e4-allele status was included in a further analysis using the different adjustment sets. All analyses were repeated and reproductive lifespan was used instead of age at menopause.

## 3 Results

Figure 2 shows a flowchart illustrating the derivation of the study populations used for the analyses after applying the global and test-specific exclusion criteria restricted to postmenopausal women. Data were available for 2,251 postmenopausal women aged 45-80 years for the Word Fluency Test, and for 2,193 postmenopausal women aged 45-80 years for TMT A and B. The characteristics of these women are shown in Table 1.

**Figure 2.**
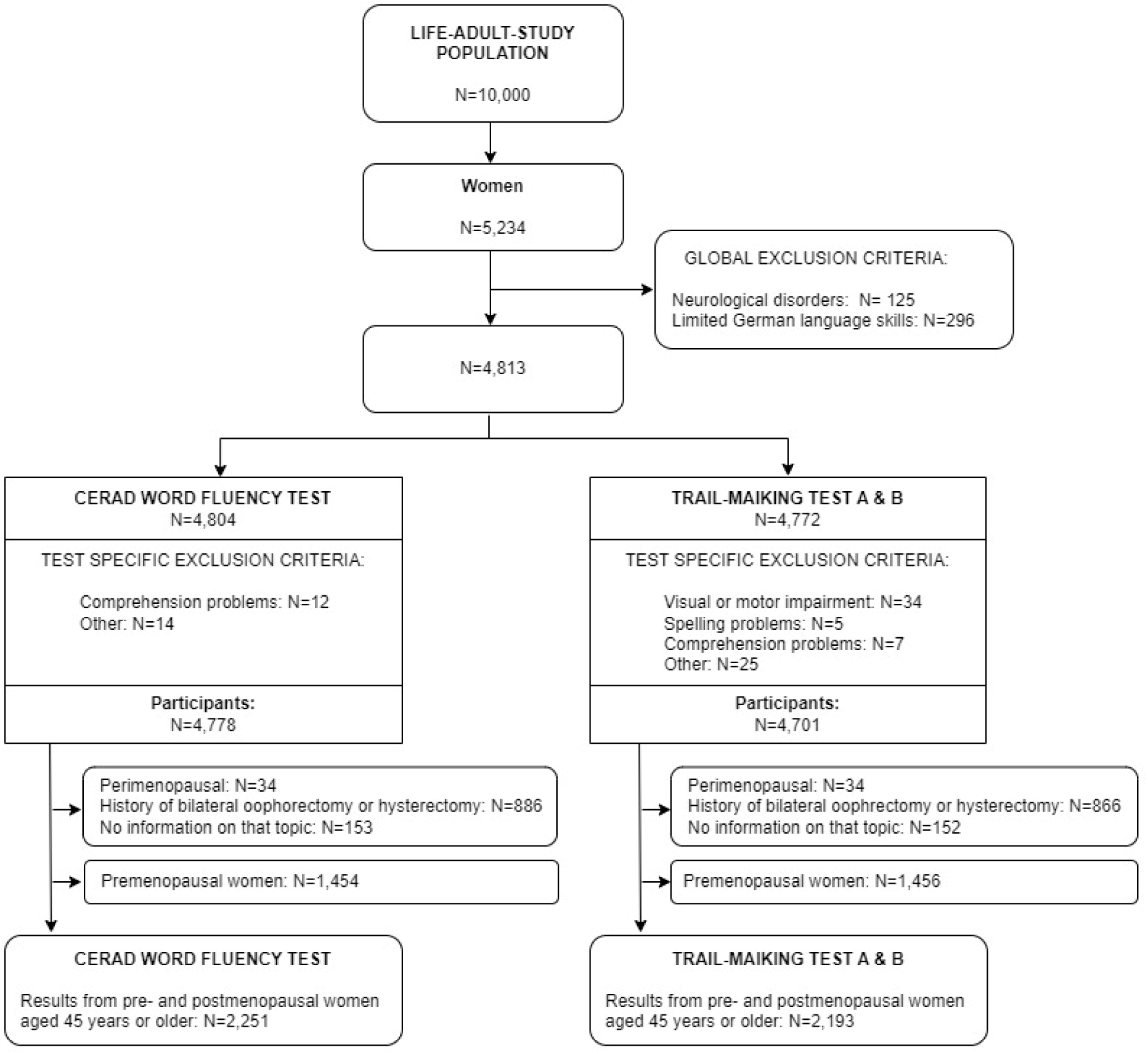
Flowchart showing the study population used in the analyses after applying global and test-specific exclusion criteria.

**Table 1:**
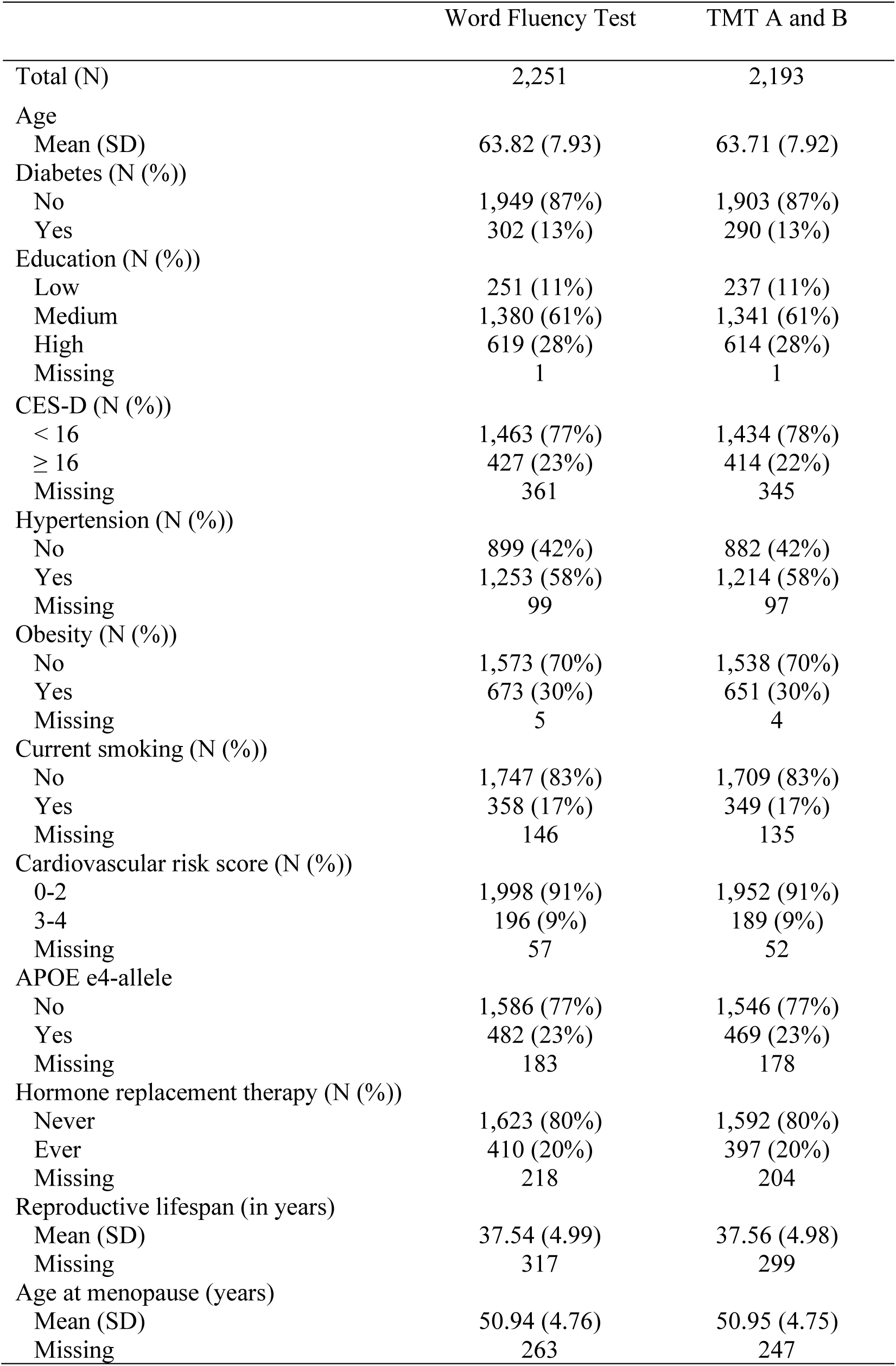
Characteristics of the LIFE-Adult study population of postmenopausal women aged 45-80 years.

The mean age was 63.7 years (Word Fluency Test) and 63.8 years (TMT A and B). Overall, 9% of the women had a cardiovascular risk score of 3-4 and 23% were carriers of at least one APOE e4 allele. Low education was reported in 11% of the women, medium education in 61% and high education in 28%. Hormone replacement therapy was used by 20% of the women at some time. The average length of the reproductive lifespan was 37.5 years.

Table 2 shows the estimates of the total effects of age at menopause (per 10 years) on cognitive performance on the Word Fluency Test, TMT A, and TMT B in postmenopausal women. Age at menopause was positively associated with all three cognitive tests analyzed using the basic adjustment (Word Fluency Test: 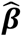 = 0.11, 95% CI = [0.01, 0.20], TMT A: 0.14 [0.05, 0.23], and TMT B: 0.12 [0.03, 0.22]). An additional adjustment for cardiovascular risk score and CES-D showed similar results, although only significant for TMT A and B.

**Table 2:** Total effect of age at menopause and reproductive lifespan on cognitive test results in postmenopausal women using different adjustment sets. The basic adjustment variables are: age, age^2^, education, and APOE e4 allele status. As a sensitivity analysis, cardiovascular risk score and CES-D are added to the basic adjustment set. All z-scores are scaled so that higher values represent better outcomes. Participants with missing values were excluded from the analysis.

| Outcome | Basic adjustment |  | Additionally adjusted for cardiovascular risk score and CES-D |  |
| --- | --- | --- | --- | --- |
| | N | $\hat{\beta}$ [95% CI] | N | $\hat{\beta}$ [95% CI] |
| **Age at menopause (10 years)** |  |  |  |  |
| Word Fluency | 1830 | 0.11 * [0.01, 0.20] | 1576 | 0.09 [-0.02, 0.20] |
| TMT A | 1791 | 0.14 ** [0.05, 0.23] | 1546 | 0.10 * [0.01, 0.20] |
| TMT B | 1791 | 0.12 ** [0.03, 0.22] | 1546 | 0.11 * [0.01, 0.21] |
| **Reproductive lifespan (10 years)** |  |  |  |  |
| Word Fluency | 1780 | 0.12 * [0.03, 0.22] | 1533 | 0.11 * [0.00, 0.21] |
| TMT A | 1743 | 0.11 * [0.02, 0.20] | 1505 | 0.07 [-0.02, 0.17] |
| TMT B | 1743 | 0.10 * [0.02, 0.19] | 1505 | 0.09 [-0.01, 0.18] |

\*\*\* p < 0.001; \*\* p < 0.01; \* p < 0.05

Significant positive associations were also observed for reproductive lifespan with all three neuropsychological tests considered when the basic adjustment was used (Word Fluency Test 0.12 [0.03, 0.22], TMT A 0.11 [0.02, 0.20] and TMT B 0.10 [0.02, 0.19]). Using the additional adjustment, there were also positive associations between reproductive lifespan and neuropsychological tests, although not always significant.

An analysis including only postmenopausal women not taking HRT also showed similar, positive associations of age at menopause and reproductive lifespan with cognitive test scores (Table 3).

**Table 3:**
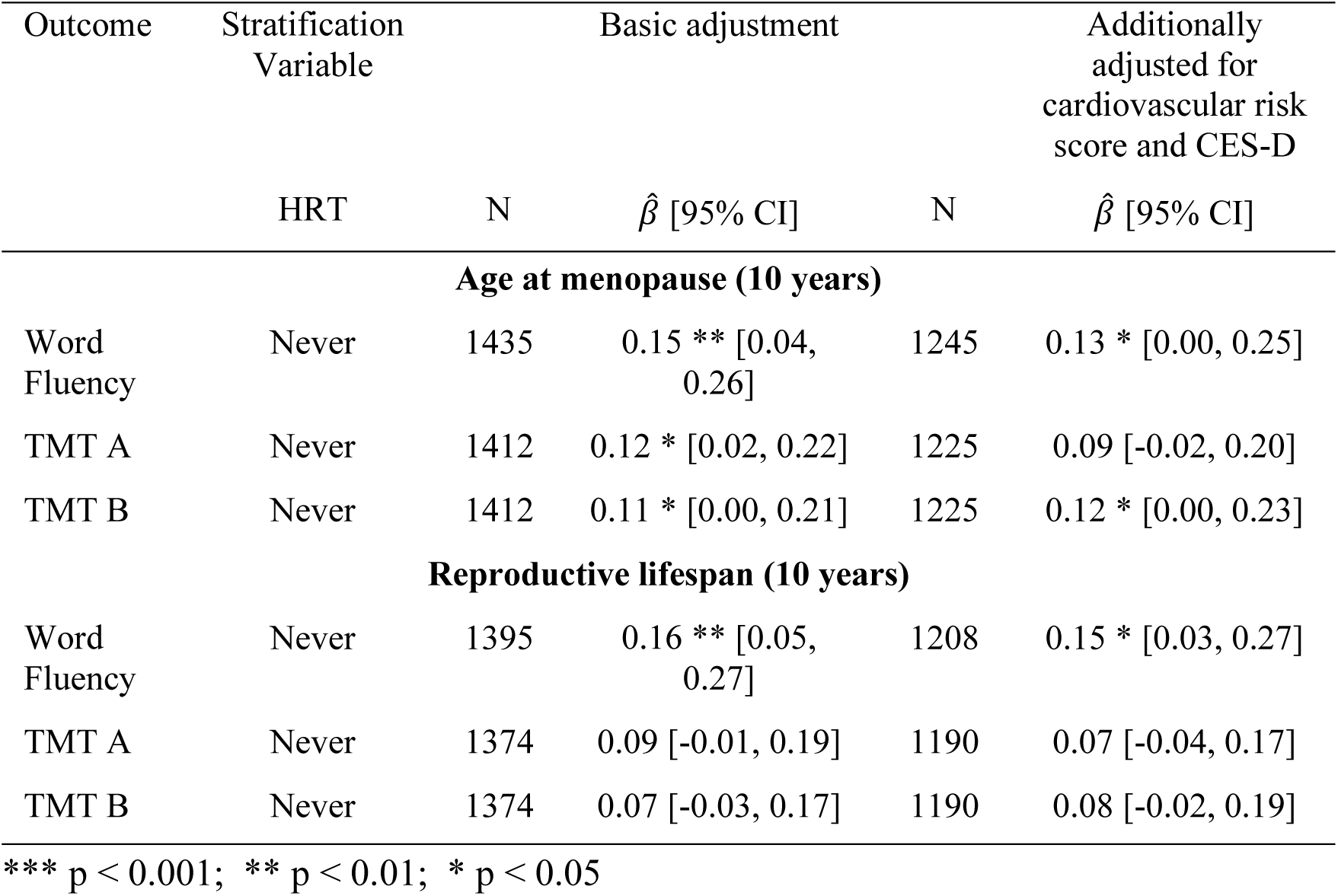
Total effect of age at menopause and reproductive lifespan on cognitive test results in postmenopausal women without prior HRT use using different adjustment sets. The basic adjustment variables are: age, age^2^, education, and APOE e4-allele status. As sensitivity analysis, cardiovascular risk score and CES-D are added to the basic adjustment set. All z- scores are scaled so that higher values represent better outcomes. Participants with missing values were excluded from the analysis.

| Outcome | Stratification Variable |  | Basic adjustment |  | Additionally adjusted for cardiovascular risk score and CES-D |
| --- | --- | --- | --- | --- | --- |
| | HRT | N | $\hat{\beta}$ [95% CI] | N | $\hat{\beta}$ [95% CI] |
| **Age at menopause (10 years)** |  |  |  |  |  |
| Word Fluency | Never | 1435 | 0.15 ** [0.04, 0.26] | 1245 | 0.13 * [0.00, 0.25] |
| TMT A | Never | 1412 | 0.12 * [0.02, 0.22] | 1225 | 0.09 [-0.02, 0.20] |
| TMT B | Never | 1412 | 0.11 * [0.00, 0.21] | 1225 | 0.12 * [0.00, 0.23] |
| **Reproductive lifespan (10 years)** |  |  |  |  |  |
| Word Fluency | Never | 1395 | 0.16 ** [0.05, 0.27] | 1208 | 0.15 * [0.03, 0.27] |
| TMT A | Never | 1374 | 0.09 [-0.01, 0.19] | 1190 | 0.07 [-0.04, 0.17] |
| TMT B | Never | 1374 | 0.07 [-0.03, 0.17] | 1190 | 0.08 [-0.02, 0.19] |

\*\*\* $p < 0.001$ ; \*\* $p < 0.01$ ; \* $p < 0.05$

We found no significant interaction between age at menopause wand APOE e4 allele status in postmenopausal women (all p-values >0.1).

## 4 Discussion

This study aimed to disentangle the effects of cumulative estrogen exposure on cognition in women using a causal framework approach. We found that a longer reproductive lifespan and a later age at menopause were associated with better cognitive performance in postmenopausal women, particularly in in the domains of processing speed and working memory.

Our findings contribute to the growing body of evidence on the relationship between reproductive factors and cognitive function in women. A comprehensive review by Georgakis et al. (2016) also suggested that later age at menopause and a longer reproductive lifespan were associated with better cognitive function in postmenopausal women [39]. Our findings are consistent with these conclusions, providing additional support for the potential cognitive benefits of prolonged exposure to endogenous estrogens. Our results are also consistent with recent findings by Lindseth et al. (2022), who examined associations between reproductive history, hormone use, and cognition in a large sample of middle-to older-aged women from the UK Biobank [12]. They also reported that a longer reproductive lifespan and older age at menopause were positively associated with cognitive performance in processing speed and working memory.

Other work in this area asks slightly different, not necessarily causal, questions and uses different exposures, respectively. For example, recent research by Than et al (2023), also using data from the UK Biobank, compared the rate of cognitive change in women who were either premenopausal, perimenopausal or postmenopausal. They found that compared with premenopausal women, perimenopausal and postmenopausal women showed a greater rate of decline in psychomotor speed, but not in fluid intelligence and memory, over an average of 8 years [40]. Epperson et al. (2013) also analyzed the cognitive change during the menopausal transition and found an effect on verbal memory performance (Buschke Selective Reminding Test), but not on the performance in other analyzed tests (Digit Symbol Substitution Test and Symbol Copy Task). Although these studies are longitudinal and raise important hypotheses, they do not ask questions using defined causal exposures. It is also important to note that menopause itself, defined as a period of time, and the menopausal transition cannot be considered as causal exposures.

In contrast, the use of Directed Acyclic Graphs (DAGs), forces explicit assumptions about the relationships between menopause-related exposures, cognitive outcomes, and potential confounders and mediators. This approach allows the identification of appropriate adjustment sets, providing a more robust basis for targeted prevention strategies.

By using this causal framework, our study adds several important dimensions to the existing evidence. First, we tried to define causal exposures and decided to focus on lifelong estrogen exposure, using age at menopause and reproductive lifespan as proxies. Second, by using DAGs we were able to more rigorously identify factors that might act as potential confounders, moderators and mediators in the relationship between age at menopause (reproductive years) and cognition. This is particularly important because some factors, such as depression or cardiovascular disease, may play different roles at different times in life.

The role of cardiovascular disease (CVD) and depression in the relationship between age of menopause and cognition is complex and warrants further discussion. It is important to note that the relationships between these variables may be bidirectional, and the timing of their onset relative to menopause may affect their role as mediators or confounders. Interestingly, recent research suggests that CVD may be a risk factor for early menopause, rather than simply a consequence of estrogen depletion [11]. Kok et al. (2006) found that cardiovascular risk factors may influence the age at menopause, highlighting the complex interplay between cardiovascular health and reproductive aging [41]. Similarly, depression may contribute to an earlier onset of menopause [42], and, conversely, age at menopause and reproductive lifespan might affect the risk of developing depression [43]. These findings highlight the importance of considering the temporal relationships between these factors in future research. In our study, we therefore considered these factors both as potential mediators (main analysis) and as potential confounders (sensitivity analysis) in the causal pathway between age at menopause and cognition.

A major strength of our study is the use of a population-based sample and the comprehensive assessment of cognitive function. However, several limitations must be acknowledged. First, the cross-sectional nature of our study limits our ability to fully apply our defined adjustments and thus to draw definitive causal conclusions. Longitudinal studies with repeated cognitive assessments before, during, and after the menopausal transition as well as time-specific information on potential mediating and moderating and confounding factors would provide stronger evidence of causal effects. To address this, sensitivity analyses were carried out using a confounder and a mediating approach. Second, although we attempted to account for potential confounding factors through our DAG approach, residual confounding may exist, particularly through factors we are not aware to act as confounders. Third, our reliance on self-reported menopausal status and age at menopause may introduce misclassification. However, we believe that non-differential misclassification would likely bias our results towards the null, potentially underestimating the true associations between reproductive factors and cognitive function.

In conclusion, our study provides evidence for associations between age at menopause and reproductive lifespan, respectively, and cognitive function in middle-aged and older women, supporting and extending the findings of, for example, Georgakis et al. (2016) [39] and Lindseth et al. (2022) [12]. The use of a causal framework approach strengthens the validity of our findings and highlights the complex interplay between menopause-related exposures, and cognition. Future research should focus on longitudinal assessments of cognitive function throughout the menopausal transition, incorporate biomarkers of hormonal status, and further explore the potential modifying effects of lifestyle factors and genetic predisposition. Such studies, combined with the causal framework approach used in our research, could provide even stronger evidence of the causal relationships between reproductive factors and cognitive function in women. This work will be crucial in developing targeted interventions to maintain cognitive health in aging women.

## Data availability statement

Data from the LIFE-Adult-Study are available to researchers who submit a detailed written proposal, including objectives, measures, names of all researchers involved, and how results and newly generated data will be returned for further use. Data are provided upon approval by the data use- and access-committee. Inquiries are to be submitted to.

## Author Contributions

NB drafted the manuscript and conducted and programmed all statistical analysis. HM came up with the research question, supervised the analyses, and supported the drafting of the manuscript. CE, MLö, MLS, AV, SRH designed the LIFE-Adult-Study. AEZ, SR, MLu, KW, CE, MLö, RB, MLS, AV, JS, VW and SRH made substantial contributions to the data acquisition and revised the manuscript critically for important intellectual content, respectively. All authors approved the final version of the manuscript to be published.

## Acknowledgments

We thank all participants of the LIFE-Adult-Study for their good collaboration as well as all members of the LIFE study center for conducting the study.

## Literature

1. What Is Menopause? National Institute on Aging. October 16, 2024. Accessed March 5, 2025. https://www.nia.nih.gov/health/menopause/what-menopause

2. Berent-Spillson A, Persad CC, Love T, et al. Hormonal Environment Affects Cognition Independent of Age during the Menopause Transition. J Clin Endocrinol Metab. 2012;97(9):E1686–E1694. doi:10.1210/jc.2012-1365

3. Epperson CN, Sammel MD, Freeman EW. Menopause effects on verbal memory: findings from a longitudinal community cohort. J Clin Endocrinol Metab. 2013;98(9):3829–3838. doi:10.1210/jc.2013-1808

4. Hogervorst E, Craig J, O’Donnell E. Cognition and mental health in menopause: A review. Best Pract Res Clin Obstet Gynaecol. 2022;81:69–84. doi:10.1016/j.bpobgyn.2021.10.009

5. Arevalo MA, Azcoitia I, Garcia-Segura LM. The neuroprotective actions of oestradiol and oestrogen receptors. Nat Rev Neurosci. 2015;16(1):17–29. doi:10.1038/nrn3856

6. Heys M, Jiang C, Cheng KK, et al. Life long endogenous estrogen exposure and later adulthood cognitive function in a population of naturally postmenopausal women from Southern China: the Guangzhou Biobank Cohort Study. Psychoneuroendocrinology. 2011;36(6):864–873. doi:10.1016/j.psyneuen.2010.11.009

7. Morrison JH, Brinton RD, Schmidt PJ, Gore AC. Estrogen, menopause, and the aging brain: how basic neuroscience can inform hormone therapy in women. J Neurosci Off J Soc Neurosci. 2006;26(41):10332–10348. doi:10.1523/JNEUROSCI.3369-06.2006

8. Fox M, Berzuini C, Knapp LA. Cumulative estrogen exposure, number of menstrual cycles, and Alzheimer’s risk in a cohort of British women. Psychoneuroendocrinology. 2013;38(12):2973–2982. doi:10.1016/j.psyneuen.2013.08.005

9. Greendale GA, Huang MH, Wight RG, et al. Effects of the menopause transition and hormone use on cognitive performance in midlife women. Neurology. 2009;72(21):1850–1857. doi:10.1212/WNL.0b013e3181a71193

10. Henderson VW, Popat RA. Effects of endogenous and exogenous estrogen exposures in midlife and late-life women on episodic memory and executive functions. Neuroscience. 2011;191:129–138. doi:10.1016/j.neuroscience.2011.05.059

11. Hemachandra C, Taylor S, Islam RM, Fooladi E, Davis SR. A systematic review and critical appraisal of menopause guidelines. BMJ Sex Reprod Health. 2024;50(2):122–138. doi:10.1136/bmjsrh-2023-202099

12. Lindseth LRS, de Lange AMG, van der Meer D, et al. Associations between reproductive history, hormone use, APOE ε4 genotype and cognition in middle- to older-aged women from the UK Biobank. Front Aging Neurosci. 2022;14:1014605. doi:10.3389/fnagi.2022.1014605

13. Pearl J. Causality: Models, Reasoning and Inference. 2nd Edition. Cambridge University Press; 2009.

14. Glymour MM, Weuve J, Chen JT. Methodological challenges in causal research on racial and ethnic patterns of cognitive trajectories: measurement, selection, and bias. Neuropsychol Rev. 2008;18(3):194–213. doi:10.1007/s11065-008-9066-x

15. Hernán MA, Robins JM. Causal Inference: What If. Boca Raton: Chapman & Hall/CRC; 2020.

16. Loeffler M, Engel C, Ahnert P, et al. The LIFE-Adult-Study: objectives and design of a population-based cohort study with 10,000 deeply phenotyped adults in Germany. BMC Public Health. 2015;15(1):691. doi:10.1186/s12889-015-1983-z

17. Engel C, Wirkner K, Zeynalova S, et al. Cohort Profile: The LIFE-Adult-Study. Int J Epidemiol. 2023;52(1):e66–e79. doi:10.1093/ije/dyac114

18. Kynast J, Quinque EM, Polyakova M, et al. Mindreading From the Eyes Declines With Aging - Evidence From 1,603 Subjects. Front Aging Neurosci. 2020;12:550416. doi:10.3389/fnagi.2020.550416

19. Luck T, Roehr S, Rodriguez FS, et al. Memory-related subjective cognitive symptoms in the adult population: prevalence and associated factors – results of the LIFE-Adult-Study. BMC Psychol. 2018;6(1):23. doi:10.1186/s40359-018-0236-1

20. Schroeter ML, Kynast J, Schlögl H, Baron-Cohen S, Villringer A. Sex and age interact in reading the mind in the eyes. Compr Psychoneuroendocrinology. 2022;12:100162. doi:10.1016/j.cpnec.2022.100162

21. Brauns H, Scherer S, Steinmann S. The CASMIN Educational Classification in International Comparative Research. In: Hoffmeyer-Zlotnik JHP, Wolf C, eds. Advances in Cross-National Comparison: A European Working Book for Demographic and Socio-Economic Variables. Springer US; 2003:221–244. doi:10.1007/978-1-4419-9186-7_11

22. Radloff LS. The CES-D Scale: A Self-Report Depression Scale for Research in the General Population. Appl Psychol Meas. 1977;1(3):385–401. doi:10.1177/014662167700100306

23. Aslanidis C, Schmitz G. High-speed apolipoprotein E genotyping and apolipoprotein B3500 mutation detection using real-time fluorescence PCR and melting curves. Clin Chem. 1999;45(7):1094–1097.

24. Reitan RM. Trail making test: manual for administration and scoring. Reitan Neuropsychology Laboratory. Published online 1992.

25. Morris JC, Mohs RC, Rogers H, Fillenbaum G, Heyman A. Consortium to establish a registry for Alzheimer’s disease (CERAD) clinical and neuropsychological assessment of Alzheimer’s disease. Psychopharmacol Bull. 1988;24(4):641–652.

26. Morris JC, Heyman A, Mohs RC, et al. The Consortium to Establish a Registry for Alzheimer’s Disease (CERAD). Part I. Clinical and neuropsychological assessment of Alzheimer’s disease. Neurology. 1989;39(9):1159–1165. doi:10.1212/wnl.39.9.1159

27. Tombaugh TN, Kozak J, Rees L. Normative data stratified by age and education for two measures of verbal fluency: FAS and animal naming. Arch Clin Neuropsychol Off J Natl Acad Neuropsychol. 1999;14(2):167–177.

28. Schmid NS, Ehrensperger MM, Berres M, Beck IR, Monsch AU. The Extension of the German CERAD Neuropsychological Assessment Battery with Tests Assessing Subcortical, Executive and Frontal Functions Improves Accuracy in Dementia Diagnosis. Dement Geriatr Cogn Disord Extra. 2014;4(2):322–334. doi:10.1159/000357774

29. Textor J, van der Zander B, Gilthorpe MS, Liśkiewicz M, Ellison GT. Robust causal inference using directed acyclic graphs: the R package ‘dagitty.’ Int J Epidemiol. 2016;45(6):1887–1894. doi:10.1093/ije/dyw341

30. Richard I, Roy O. DiagrammeR: Graph/Network Visualization, R package version 1.0.11, <https://CRAN.R-project.org/package=DiagrammeR>. Published online 2024.

31. Hare DL, Toukhsati SR, Johansson P, Jaarsma T. Depression and cardiovascular disease: a clinical review. Eur Heart J. Published online November 26, 2013:eht462. doi:10.1093/eurheartj/eht462

32. Koochmeshgi J, Hosseini-Mazinani SM, Morteza Seifati S, Hosein-Pur-Nobari N, Teimoori-Toolabi L. Apolipoprotein E genotype and age at menopause. Ann N Y Acad Sci. 2004;1019:564–567. doi:10.1196/annals.1297.105

33. Meng FT, Wang YL, Liu J, Zhao J, Liu RY, Zhou JN. ApoE genotypes are associated with age at natural menopause in Chinese females. Age Dordr Neth. 2012;34(4):1023–1032. doi:10.1007/s11357-011-9287-4

34. Pontifex MG, Martinsen A, Saleh RNM, et al. APOE4 genotype exacerbates the impact of menopause on cognition and synaptic plasticity in APOE-TR mice. FASEB J. 2021;35(5):e21583. doi:10.1096/fj.202002621RR

35. Smith CA, McCleary CA, Murdock GA, et al. Lifelong estrogen exposure and cognitive performance in elderly women. Brain Cogn. 1999;39(3):203–218. doi:10.1006/brcg.1999.1078

36. Rasgon NL, Magnusson C, Johansson ALV, Pedersen NL, Elman S, Gatz M. Endogenous and exogenous hormone exposure and risk of cognitive impairment in Swedish twins: a preliminary study. Psychoneuroendocrinology. 2005;30(6):558–567. doi:10.1016/j.psyneuen.2005.01.004

37. R Core Team. R: A Language and Environment for Statistical Computing. Published online 2023. https://www.R-project.org/

38. RStudio Team. RStudio: Integrated Development Environment for R. Published online 2023. http://www.rstudio.com/

39. Georgakis MK, Kalogirou EI, Diamantaras AA, et al. Age at menopause and duration of reproductive period in association with dementia and cognitive function: A systematic review and meta-analysis. Psychoneuroendocrinology. 2016;73:224–243. doi:10.1016/j.psyneuen.2016.08.003

40. Than S, Moran C, Beare R, et al. Cognitive trajectories during the menopausal transition. Front Dement. 2023;2:1098693. doi:10.3389/frdem.2023.1098693

41. Kok HS, van Asselt KM, van der Schouw YT, et al. Heart disease risk determines menopausal age rather than the reverse. J Am Coll Cardiol. 2006;47(10):1976–1983. doi:10.1016/j.jacc.2005.12.066

42. Harlow BL, Signorello LB. Factors associated with early menopause. Maturitas. 2000;35(1):3–9. doi:10.1016/S0378-5122(00)00092-X

43. Wu Q, Yan Y, La R, et al. Association of reproductive lifespan and age at menopause with depression: Data from NHANES 2005-2018. J Affect Disord. 2024;356:519–527. doi:10.1016/j.jad.2024.04.077

