## Supplemental Material for "Disentangling the effects of menopause on cognitive performance – causal framework and findings of the LIFE-Adult-Study"

**Supplemetal Table 1:** The directed acyclic graph (DAG) illustrates the complex relationships between age at menopause and reproductive span, respectively, (the exposure) and cognition (the outcome), along with various other factors that may influence this relationship. Here are the results of a literature search that established these relationships.

| **Reference** | **Main result / Quote** | **X causes Y** |
| --- | --- | --- |
| Georgakis et al. (2016) [1] | Increasing age at menopause and reproductive period duration are possibly associated with better cognitive performance and delayed cognitive decline. | Age at menopause 🡪 Cognition  Reproductive period 🡪 Cognition |
| Lindseth et al. (2022) [2] | A longer reproductive span, older age at menopause, older age at first and last birth, and use of hormonal contraceptives were positively associated with cognitive performance later in life.  While APOE ε4 genotype was associated with reduced processing speed and executive functioning, in a dose-dependent manner, it did not influence the observed associations between female-specific factors and cognition. | Reproductive period 🡪 Cognition  Age at menopause 🡪 Cognition  Number of live births 🡪 Cognition (Task specific)  HRT 🡪 Cognition (Task specific)  APOE ε4 genotype 🡪 Cognition |
| Wu et al. (2024) [3] | They suggested that a shorter reproductive lifespan and an earlier age at menopause were associated with the development of depression. | Age at menopause 🡪 Depression  Reproductive lifespan 🡪 Depression |
| Krittanawong et al. (2023) [4] | Depression has a significant negative impact on development of cardiovascular disease and on cardiovascular disease outcomes | Depression 🡪 Cardiovascular disease |
| Hare et al. (2013) [5] | Patients with CVD have more depression than the general population. Persons with depression are more likely to eventually develop CVD. Given the increased prevalence of depression in patients with CVD, a causal relationship with either CVD causing more depression or depression causing more CVD and a worse prognosis for CVD is probable. | Depression 🡪 Cardiovascular disease  Cardiovascular disease 🡪 Depression |
| Harlow et al. (1995) [6] | They suggested a link between a self-reported history of medically treated depression and early menopause. | Depression 🡪 Age at menopause |
| Kim et al. (2019) [7] | Korean women who perceived themselves to be overweight or obese, who had depressive symptoms, or who smoked or were current smokers had higher probabilities of experiencing the onset of menopause, whereas those who had educational achievement lower than high school had a lower probability of experiencing the onset of menopause. | Smoking 🡪 Age at menopause  Overweight/obese 🡪 Age at menopause  Depression 🡪 Age at menopause  Education 🡪 Age at menopause |
| Stern (2009) [8] | Higher education is associated with better cognitive function and may provide cognitive reserve | Education 🡪 Cognition |
| Liu et al. (2013) [9] | APOE genotype is a known risk factor for cognitive decline and Alzheimer's disease | APOE genotype 🡪 Cognition |
| Lyu and Burr (2016) [10] | Childhood Socioeconomic Status(SES) and adult SES both had relationships with cognitive status and, to a lesser degree, change in cognition in later life. | SES (Socioeconomic Status) 🡪 Cognition |
| Gorelick et al. (2011) [11] | Cardiovascular disease (CVD) can negatively impact cognitive function. | CVD (Cardiovascular Disease) 🡪 Cognition |
| Maki (2013) [12] | Observational data suggest that HT reduces the risk of Alzheimer’s disease (AD) generally. | HRT 🡪 Alzheimer’s disease |
| Yaffe et al. (2000) [13] | Estrogen use was associated with less cognitive decline among ε4-negative women but not ε4-positive women. Potential mechanisms, including carotid atherosclerosis, by which ε4 may interact with estrogen and cognition warrant further investigation. | Estrogen use 🡪 Cognitive decline  Only in APOE ε4 positive women |
| Rock et al. (2014) [14] | Depression can negatively impact cognitive function | Depression 🡪 Cognition |
| Greendale et al. (2009) [15] | This paper discusses the temporary decrement in cognitive performance during perimenopause and the effect of hormone initiation on cognitive performance. | Menopausal transition 🡪 Cognition  HRT 🡪 Cognition |
| Sirin et al. (2005) [16] | This meta-analytic review demonstrates the relationship between socioeconomic status and academic achievement. | SES 🡪 Education |
| Schultz et al. (2018) [17] | They reviewed the impact of socioeconomic status on cardiovascular disease risk, showing that lower SES is associated with higher CVD risk. | SES 🡪 CVD risk |
| Meng et al. (2012) [18] | This study suggests that APOE genotype may influence the age of natural menopause in Chinese females. | APOE 🡪 Age at menopause |
| Koochmeshgi et al. (2004) [19] | They found a significant relationship between APOE genotype and age at menopause. Carriers of the APOE4 allele reached menopause at an earlier age. | APOE 🡪 Age at menopause |
| Palmer et al. (2023) [20] | They reviewed new evidence regarding the impact of APOE ε4 on cognition among healthy older adults. | APOE ε4 🡪 Cognition |
| Eichner et al. (2002) [[21]](https://doi.org/10.1093/aje/155.6.487) | This review examines the association between the apolipoprotein (apo) var epsilon gene polymorphism (or its protein product (apo E)) and cardiovascular disease, among other things. | APOE 🡪 CVD risk |
| Salthouse et al. (2019) [22] | Normal cognitive aging is characterized by nearly linear declines from early adulthood in speed, and accelerating declines in memory and reasoning. However, vocabulary knowledge increased until the decade of the 60’s. | Age 🡪 Cognition |
| Toffoletto et al. (2014) [23] | There is mounting evidence attesting the relevance of endogenous ovarian hormones as well as exogenous estradiol and progesterone for emotional and cognitive processing. | Estradiol 🡪 Cognition |
| Russell et al. (2019) [24] | There are 3 common physiological estrogens, of which estradiol (E2) is seen to decline rapidly over the menopausal transition. This decline in E2 has been associated with a number of changes in the brain, including cognitive changes, effects on sleep, and effects on mood. | Estradiol 🡪 Cognition |
| Hemachandra et al. (2024) [25] | Acknowledged potential long-term menopause consequences were urogenital atrophy, and increased risks of cardiovascular disease and osteoporosis | Menopause 🡪 CVD |

**Literature**

1.	Georgakis MK, Kalogirou EI, Diamantaras AA, Daskalopoulou SS, Munro CA, Lyketsos CG, Skalkidou A, Petridou ET. Age at menopause and duration of reproductive period in association with dementia and cognitive function: A systematic review and meta-analysis. Psychoneuroendocrinology. 2016 Nov;73:224–43.

2.	Lindseth LRS, de Lange AMG, van der Meer D, Agartz I, Westlye LT, Tamnes CK, Barth C. Associations between reproductive history, hormone use, APOE ε4 genotype and cognition in middle- to older-aged women from the UK Biobank. Front Aging Neurosci. 2022;14:1014605.

3.	Wu Q, Yan Y, La R, Zhang X, Lu L, Xie R, Xue Y, Lin C, Xu W, Xu J, Huang L. Association of reproductive lifespan and age at menopause with depression: Data from NHANES 2005-2018. J Affect Disord. 2024 Jul 1;356:519–27.

4.	Krittanawong C, Maitra NS, Qadeer YK, Wang Z, Fogg S, Storch EA, Celano CM, Huffman JC, Jha M, Charney DS, Lavie CJ. Association of Depression and Cardiovascular Disease. Am J Med. 2023 Sep;136(9):881–95.

5.	Hare DL, Toukhsati SR, Johansson P, Jaarsma T. Depression and cardiovascular disease: a clinical review. Eur Heart J. 2013 Nov 26;eht462.

6.	Harlow BL, Cramer DW, Annis KM. Association of medically treated depression and age at natural menopause. Am J Epidemiol. 1995 Jun 15;141(12):1170–6.

7.	Kim YT, Cha C, Lee MR. Factors related to age at menopause among Korean women: the Korean Longitudinal Survey of Women and Families. Menopause N Y N. 2019 May;26(5):492–8.

8.	Stern Y. Cognitive reserve. Neuropsychologia. 2009 Aug;47(10):2015–28.

9.	Liu CC, Kanekiyo T, Xu H, Bu G. Apolipoprotein E and Alzheimer disease: risk, mechanisms, and therapy. Nat Rev Neurol. 2013 Feb;9(2):106–18.

10.	Lyu J, Burr JA. Socioeconomic Status Across the Life Course and Cognitive Function Among Older Adults: An Examination of the Latency, Pathways, and Accumulation Hypotheses. J Aging Health. 2016 Feb;28(1):40–67.

11.	Gorelick PB, Scuteri A, Black SE, Decarli C, Greenberg SM, Iadecola C, Launer LJ, Laurent S, Lopez OL, Nyenhuis D, Petersen RC, Schneider JA, Tzourio C, Arnett DK, Bennett DA, Chui HC, Higashida RT, Lindquist R, Nilsson PM, Roman GC, Sellke FW, Seshadri S, American Heart Association Stroke Council, Council on Epidemiology and Prevention, Council on Cardiovascular Nursing, Council on Cardiovascular Radiology and Intervention, and Council on Cardiovascular Surgery and Anesthesia. Vascular contributions to cognitive impairment and dementia: a statement for healthcare professionals from the american heart association/american stroke association. Stroke. 2011 Sep;42(9):2672–713.

12.	Maki PM. Critical window hypothesis of hormone therapy and cognition: a scientific update on clinical studies. Menopause N Y N. 2013 Jun;20(6):695–709.

13.	Yaffe K, Haan M, Byers A, Tangen C, Kuller L. Estrogen use, APOE, and cognitive decline: evidence of gene-environment interaction. Neurology. 2000 May 23;54(10):1949–54.

14.	Rock PL, Roiser JP, Riedel WJ, Blackwell AD. Cognitive impairment in depression: a systematic review and meta-analysis. Psychol Med. 2014 Jul;44(10):2029–40.

15.	Greendale GA, Huang MH, Wight RG, Seeman T, Luetters C, Avis NE, Johnston J, Karlamangla AS. Effects of the menopause transition and hormone use on cognitive performance in midlife women. Neurology. 2009 May 26;72(21):1850–7.

16.	Sirin SR. Socioeconomic Status and Academic Achievement: A Meta-Analytic Review of Research. Rev Educ Res. 2005 Sep 1;75(3):417–53.

17.	Schultz WM, Kelli HM, Lisko JC, Varghese T, Shen J, Sandesara P, Quyyumi AA, Taylor HA, Gulati M, Harold JG, Mieres JH, Ferdinand KC, Mensah GA, Sperling LS. Socioeconomic Status and Cardiovascular Outcomes. Circulation. 2018 May 15;137(20):2166–78.

18.	Meng FT, Wang YL, Liu J, Zhao J, Liu RY, Zhou JN. ApoE genotypes are associated with age at natural menopause in Chinese females. Age Dordr Neth. 2012 Aug;34(4):1023–32.

19.	Koochmeshgi J, Hosseini-Mazinani SM, Morteza Seifati S, Hosein-Pur-Nobari N, Teimoori-Toolabi L. Apolipoprotein E genotype and age at menopause. Ann N Y Acad Sci. 2004 Jun;1019:564–7.

20.	Palmer JM, Huentelman M, Ryan L. More than just risk for Alzheimer’s disease: *APOE* ε4’s impact on the aging brain. Trends Neurosci. 2023 Sep 1;46(9):750–63.

21.	Eichner JE, Dunn ST, Perveen G, Thompson DM, Stewart KE, Stroehla BC. Apolipoprotein E polymorphism and cardiovascular disease: a HuGE review. Am J Epidemiol. 2002 Mar 15;155(6):487–95.

22.	Salthouse TA. Trajectories of normal cognitive aging. Psychol Aging. 2019 Feb;34(1):17–24.

23.	Toffoletto S, Lanzenberger R, Gingnell M, Sundström-Poromaa I, Comasco E. Emotional and cognitive functional imaging of estrogen and progesterone effects in the female human brain: a systematic review. Psychoneuroendocrinology. 2014 Dec;50:28–52.

24.	Russell JK, Jones CK, Newhouse PA. The Role of Estrogen in Brain and Cognitive Aging. Neurother J Am Soc Exp Neurother. 2019 Jul;16(3):649–65.

25.	Hemachandra C, Taylor S, Islam RM, Fooladi E, Davis SR. A systematic review and critical appraisal of menopause guidelines. BMJ Sex Reprod Health. 2024 Apr 11;50(2):122–38.
